# A Literature Review and Pooled Case Analysis of Cardiofaciocutaneous Syndrome to Estimate Cancer Risk

**DOI:** 10.1101/2024.08.09.24311751

**Authors:** Jazmyn Bess, Toniya Brown, Sonia Bhala, Anaqa Faizer, Muzzammil Ahmadzada, Alicia A Livinski, Alex Pemov, Jung Kim, Philip S Rosenberg, Gina M Ney, Douglas R Stewart

## Abstract

**Background:** Cardiofaciocutaneous syndrome (CFC) is a rare disorder with multiple congenital anomalies including macrocephaly, failure to thrive, and neurocognitive delay. CFC is part “RASopathy” syndromes caused by pathogenic germline variants in *BRAF, KRAS, MAP2K1,* and *MAP2K2.* To estimate cancer risk in CFC we conducted a systematic review using case reports and series.

**Methods:** We reviewed articles and abstracted CFC cases to form a retrospective cohort based on PRISMA guidelines. Genotype-pphenotype (cancer) correlations, standardized incidence ratios (SIR), cumulative incidence and cause-specific hazard rates for cancer and cancer-free in CFC were calculated.

**Results:** This study includes 198 publications reporting 690 patients. Only 1.6% (11) had cancer, including acute lymphoblastic leukemia (ALL). Six cancer patients harbored pathogenic variants within *BRAF, MAP2K1*, and *MAP2K2*. Cumulative incidence by age 10 was 5% for cancer or cancer-free death. Hazard Ratio (death) was 1-2% until age 3 and declined thereafter. Significant SIRs were found for all sites (SIR=4.96) and ALL (SIR=24.23).

**Conclusions:** This is the largest investigation of cancer in CFC to date. Cancer risk in the CFC population is elevated but appears limited to earlier childhood. Modest case and cancer numbers could pose limitations to accurately assess cancer risk in CFC and more studies are needed.

**Systematic Review Registration:** The review was registered using PROSPERO under the identification tag CRD42023405823 (https://www.crd.york.ac.uk/prospero/display_record.php?RecordID=405823)

## Introduction

Cardiofaciocutaneous syndrome (CFC) is a rare disorder first described in 1986 and is characterized by multiple congenital anomalies including heart defects, distinctive craniofacial anomalies (including macrocephaly and bi-temporal narrowing), and cutaneous abnormalities. Most individuals also have some form of intellectual disability.^1,2^ The overall prevalence of CFC was last reported in 2012 as 1:810,000,^3^ but current RASopathy studies are seeking to better define and update this prevalence.^4^ CFC arises due to a pathogenic germline variant in *BRAF, KRAS, MAP2K1,* or *MAP2K2,* and is part of a larger group of syndromes termed RASopathies. RASopathies are a set of clinical syndromes with phenotypic overlap which result from pathogenic variants in genes involved in the RAS/Mitogen Activated Protein Kinase (MAPK) pathway signaling.^5^

The RAS/MAPK pathway is integral to normal development and is involved in nearly every aspect of a cell’s activity. Dysregulation of this pathway, therefore, is critical to disease development. Illustratively, somatic variants in RAS/MAPK genes result in cellular dysfunction and are among the most common oncogenes in all human cancer.^5^ It is therefore plausible that individuals with pathogenic germline variants in a RAS/MAPK gene, such as individuals with CFC, are also at increased risk of cancer.^6–12^ Thus, estimation of cancer risk is integral to understanding disease processes and risk management for this population.^2^ However, due to the rarity of many of the RASopathies, genotype-cancer correlates have been difficult to calculate and interpret.

A previous literature review was conducted on 226 reported CFC cases with calculations of cumulative incidence and hazard rates of cancer and cancer-free death in 2011.^12^ Because genetic testing is more readily available and technologies have improved since previous report, we sought to update and more accurately quantify cancer risk and incidence in CFC in this systematic literature review and pooled case analysis. This was completed by creating a retrospective cohort of CFC cases through systematic evidence review (SER) of the pertinent global biomedical literature. These cases were analyzed by determining the following: 1) a description of the types and frequencies of cancer; 2) a list of *BRAF, KRAS, MAP2K1* and *MAP2K2* pathogenic/likely pathogenic variants detected in patients when available; 3) genotype-phenotype (cancer) correlations; and 4) a pooled analysis of individual case data with estimations of cumulative incidence and hazard rates for cancer and death, and standardized incidence ratios for cancer.

## Materials and Methods

### Systematic Literature Review

We followed the PRISMA system, including the checklist and flowchart. The review was registered using PROSPERO under the identification tag CRD42023405823 (https://www.crd.york.ac.uk/prospero/display_record.php?RecordID=405823) A biomedical librarian from the National Institutes of Health library assisted with identifying articles from Embase, LILACS, PubMed, Scopus, and Web of Science databases between 1986 and Feb 2023. Although CFC was first named and identified in 1986, the timeline was extended to include any potential retroactively identified cases included in literature. The diagnostic criterion set by Reynolds and co-authors;^1^ was used to include/exclude any cases identified. We searched published qualitative, quantitative, and mixed methods studies, however “grey literature” (i.e. dissertations and conference abstracts/proceedings) was not included **(Supplementary Figure 1)**. Information regarding databases, search strategies, study selection, details of full text review, data extraction, risk of bias,^13,14^, and statistical analysis is shown in the **Supplementary Methods**.

### Study Selection

Eligibility inclusion included individuals with a clinical and/or molecular diagnosis of CFC with or without cancer phenotype from a study. Type of study designs included were case reports, case series, and cohort studies. A total of 22 studies in languages other than English were excluded. Title and abstract screening as well as full text review were done using the Covidence systematic review software tool (https://www.covidence.org/) (Veritas Health Innovation, Melbourne, Australia). A pilot of both screening steps was conducted with all reviewers on a sample of the same 15 records. Title and abstract screening were performed by four reviewers independently (TB, JB, SB, MA). Full text review was proceeded by four reviewers for articles that were deemed relevant for review and analysis (TB, JB, SB, MA). All conflicts at either stage were resolved by consensus discussion. If a conflict was not resolved, a fourth reviewer (GN) rendered a final decision.

### Data extraction

Three reviewers (TB, JB, AF) extracted variables using Google Forms. Formatting of variables and reporting was taken from Astiazaran-Symonds et al., 2023.^15^ If cases appeared in multiple articles, we only included the cases once from the source with the most detailed phenotypic data and/or longest follow-up. All cases reported in a study were extracted for variables including: 1) sex/gender; 2) ethnicity/race/nationality; 3) age at publication; 4) age at diagnosis of CFC; 5) molecular diagnosis (specific variant when provided); 6) type of cancer reported; 7) age at cancer diagnosis; 8) country of origin for cancer cases; 9) dead or alive; 10) age at death; and 11) last known age alive.

### Ethnicity classification

Race and ethnicity categories of CFC patients were classified using the Office of Management and Budget (OMB) data collection standards criteria^16^: Asian, Black or African American or African descent, White, and More than one race. An ‘Unknown’ race category was created to include Hispanic ethnicity cases and cases where race was not reported. Ethnicity was classified as Hispanic or Latino, Non-Hispanic or Latino, or unknown. Further details for race consolidation can be found in **Supplementary Methods.**

### Variant classification

If available, we collected variant information reported for *BRAF*, *MAP2K1*, *MAP2K2*, and *KRAS*. All variants were run through the Automated Germline Variant Pathogenicity (AutoGVP) to confirm and retrieve most updated ClinVar classification.^19^ For variant nomenclature and classification, we used MANE select transcripts https://www.ncbi.nlm.nih.gov/refseq/MANE/ NM_002755.3 (*MAP2K1*), NM_004333.6 (*BRAF*), NM_004985.5 (*KRAS*), and NM_030662.3 (*MAP2K2*) **(Supplementary Table 1)**. Collected cases were categorized into two binning tiers for sensitivity analysis as follows: tier 1) P/LP variants in ClinVar and/or InterVar; tier 2) cases with variants that are classified as variant of unknown significance (VUS), likely benign, large gene deletion, variant typographical errors, and cases with only a clinical diagnosis (i.e., no variant data provided).

### Statistical Analyses

The total number of cases available for qualitative analyses was 690. A subset of cases was used for sensitivity analysis that included only tier 1 criteria (pathogenic/likely variants). Cumulative incidence curves and cause-specific hazards were calculated, for overall cohort and for sensitivity analysis, to examine competing risks for cancer and death with outcomes of interest being cancer or death^12^.

Genotype-phenotype (cancer) associations were not calculated for the overall cohort due to an insufficient number of cases for analysis. However, we sought to assess genotype-phenotype (cancer) correlations found in the cohort by reporting frequency and percentages of cancers and variants associated with cancer types.

Standard incidence ratio calculations were performed using SAS v9.4. Observed cancer incidence and person-time at risk were derived from literature reports, assuming that each person was at risk from birth until publication date and including all reported cancers excluding non-melanoma skin cancer (which is not available in the expected rates). Population cancer incidence rates were created using SEER*Stat 8.4.3 based on cancer registry data from 8 registries for 1975-2020.^20^ Expected values for the calculations were calculated as a time-dependent weighted sum of person-time and population cancer incidence rates, stratified by sex, attained age, and attained calendar year.

## Results

### Study cohort characteristics

From the literature search, a total of 3,199 articles were reviewed and 3,001 were excluded. From the remaining 198 literature reports, a total of 690 individual cases with a clinical and/or molecular diagnosis of CFC met inclusion criteria **(Supplementary Table 2)**. Of these 690 cases, 540 reported a molecular diagnosis. **Supplementary Table 3** outlines different subsets of cases used in each analysis.

Race and ethnicity reported for CFC cases in the literature were limited, making accurate assessment of patient demographics difficult to analyze in the SER. As a result, race, ethnicity, and other demographic variables were only used for a descriptive analysis as presented in **Supplementary Table 4.** Country of origin was noted for cancer cases only, with majority of cancer cases reported from the United States and Japan **(Table 1 and Supplementary Table 2)**.

**Table 1.**
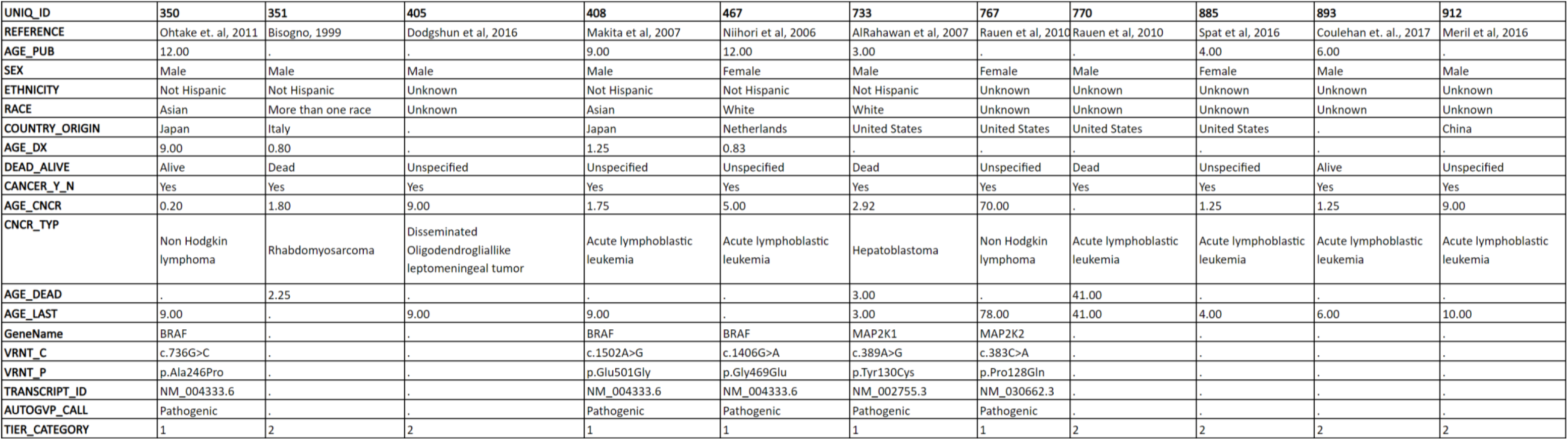
Eleven CFC cases with cancer, demographics, age, genetic variants, and tier category.

Cardiofaciocutaenous syndrome can be diagnosed both through clinical evaluation based on phenotypic features and/or through genetic testing. Of the 540 individuals with reported gene variants, there were 405 individuals with *BRAF*, 65 with *MAP2K1*, 46 with *MAP2K2*, and 24 harboring *KRAS*. A total of 101 unique amino acid changes were identified; 57 within *BRAF*, 16 within *MAP2K1*, 18 within *MAP2K2*, and 10 within *KRAS*. In total, there were 477 individuals with pathogenic variants, 33 with likely pathogenic variants, 24 with a VUS, 1 with a likely benign variant, and 5 with variant typos/discrepancies where accurate variant information could not be determined **(Figure 1A-D)**. Frequencies and percentages of the most common pathogenic/likely pathogenic variants in each gene are outlined in **Supplementary Table 5.** The most common pathogenic variants for each gene were *BRAF*: p.Gln257Arg, p. Gly469Glu, and p.Asn581Asp; *MAP2K1*: p.Tyr130Cys, p.Gly128Val; *MAP2K2*: p.Pro128Gln, p.Phe57Cys, and p.Lys61Glu; and *KRAS*: p.Gly60Arg, p.Asp153Val, and p.Pro34Arg. Additionally, there were 17 CFC cases with gene deletions, the majority of which occurred in *MAP2K2* (n=8), followed by *BRAF* (n=2). The seven remaining cases of CFC were reported as deletions and/or duplications without a specified gene.

**Figure 1.**
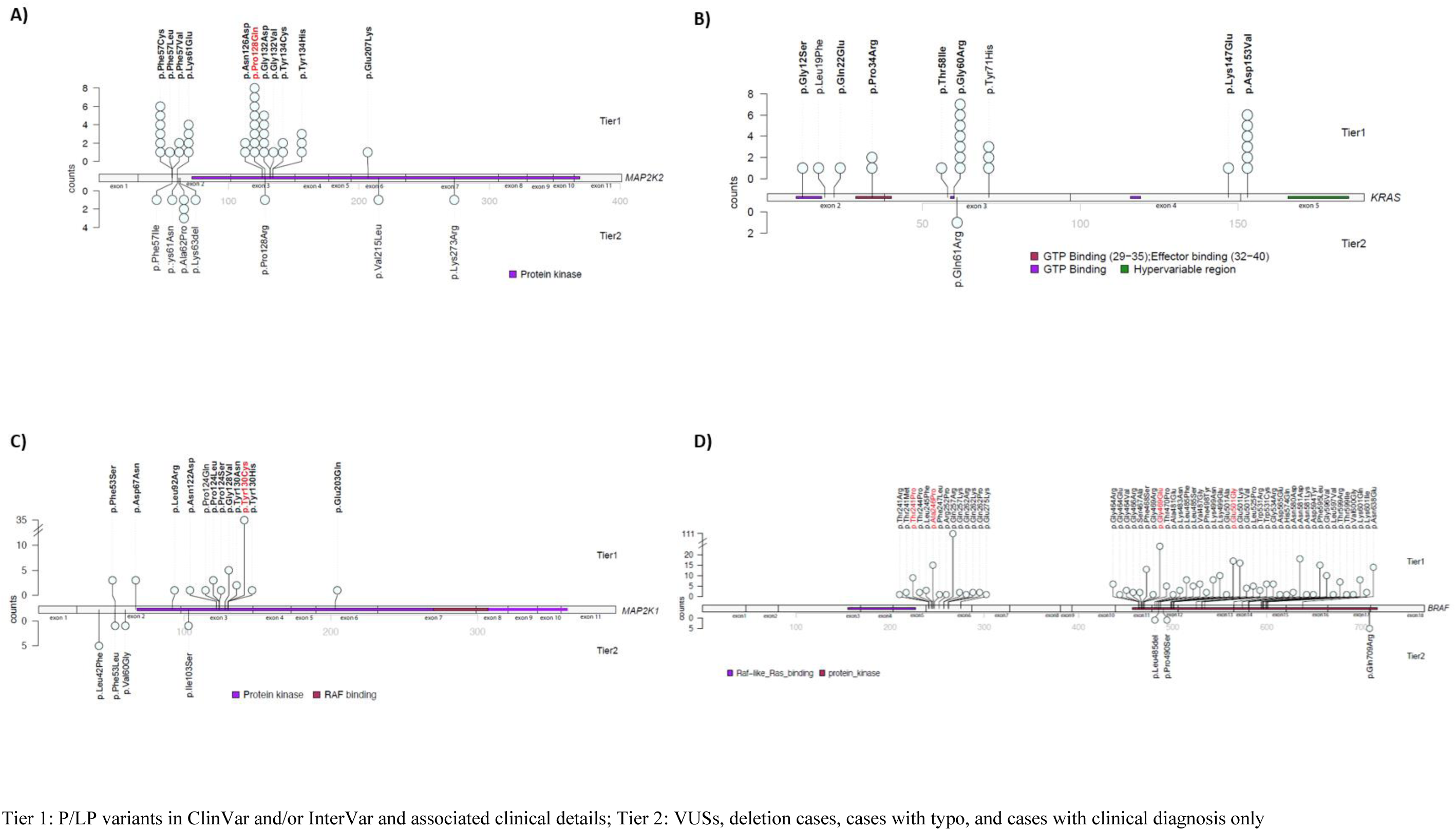
(A-D). Variants in the A) *MAP2K2,* B) *KRAS,* C) *MAP2K1*, and D) *BRAF* genes found in patients with CFC syndrome

### Genotype-phenotype (cancer) correlations

Cancer cases extracted with a clinical and/or molecular diagnosis of CFC are provided in **Table 1**. Of the 690 cases, a total of 11 cancer cases were reported, similar to a previous report count^12^. The most common cancers reported were acute lymphoblastic leukemia (ALL) (n=6, 0.87%) and non-Hodgkin lymphoma (n=2, 0.29%) **(Figure 2 and Table 2).** Single cancer cases included hepatoblastoma, rhabdomyosarcoma, and disseminated oligodendrogliallike leptomeningeal tumor (0.15%). No individuals were reported with more than one primary cancer. Six of the 11 cancers reported genetic variants, all pathogenic, within *BRAF, MAP2K1*, and *MAP2K2.* One of the most common *MAP2K2* pathogenic variants, p.Pro128Gln (25%), in the CFC cohort was reported in two cancers: ALL and non-Hodgkin lymphoma. Three of the six ALL cases reported pathogenic variants: p.Glu501Gly and p.Gly469Glu (found in the cohort at 6.1%) in *BRAF* and p.Pro128Gln in *MAP2K2*. A common *MAP2K1* pathogenic variant, p.Tyr130Cys (61.4%), was also found in the hepatoblastoma case.

**Figure 2.**
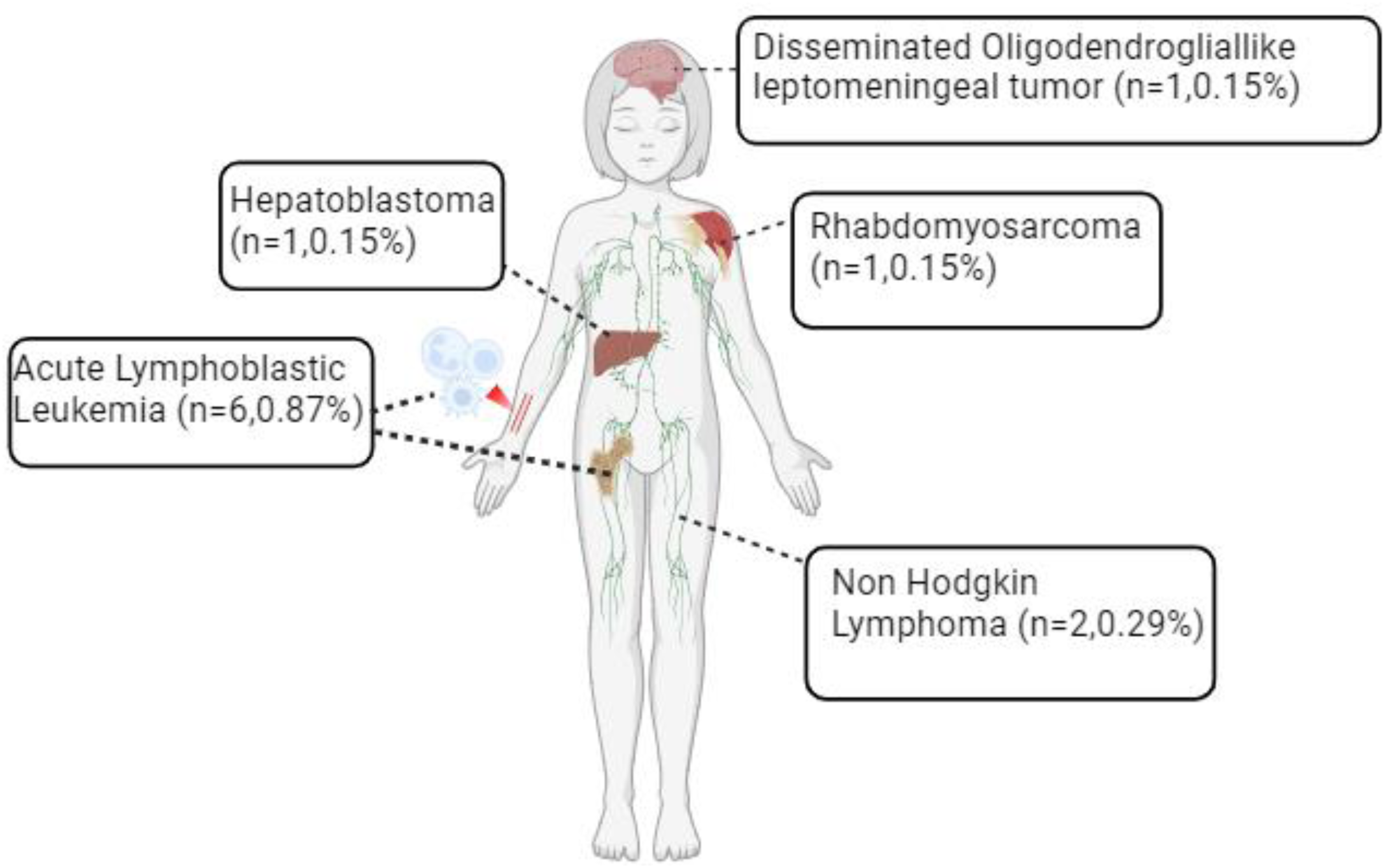
Types of cancer reported in 11 cases with CFC Syndrome

**Table 2.**
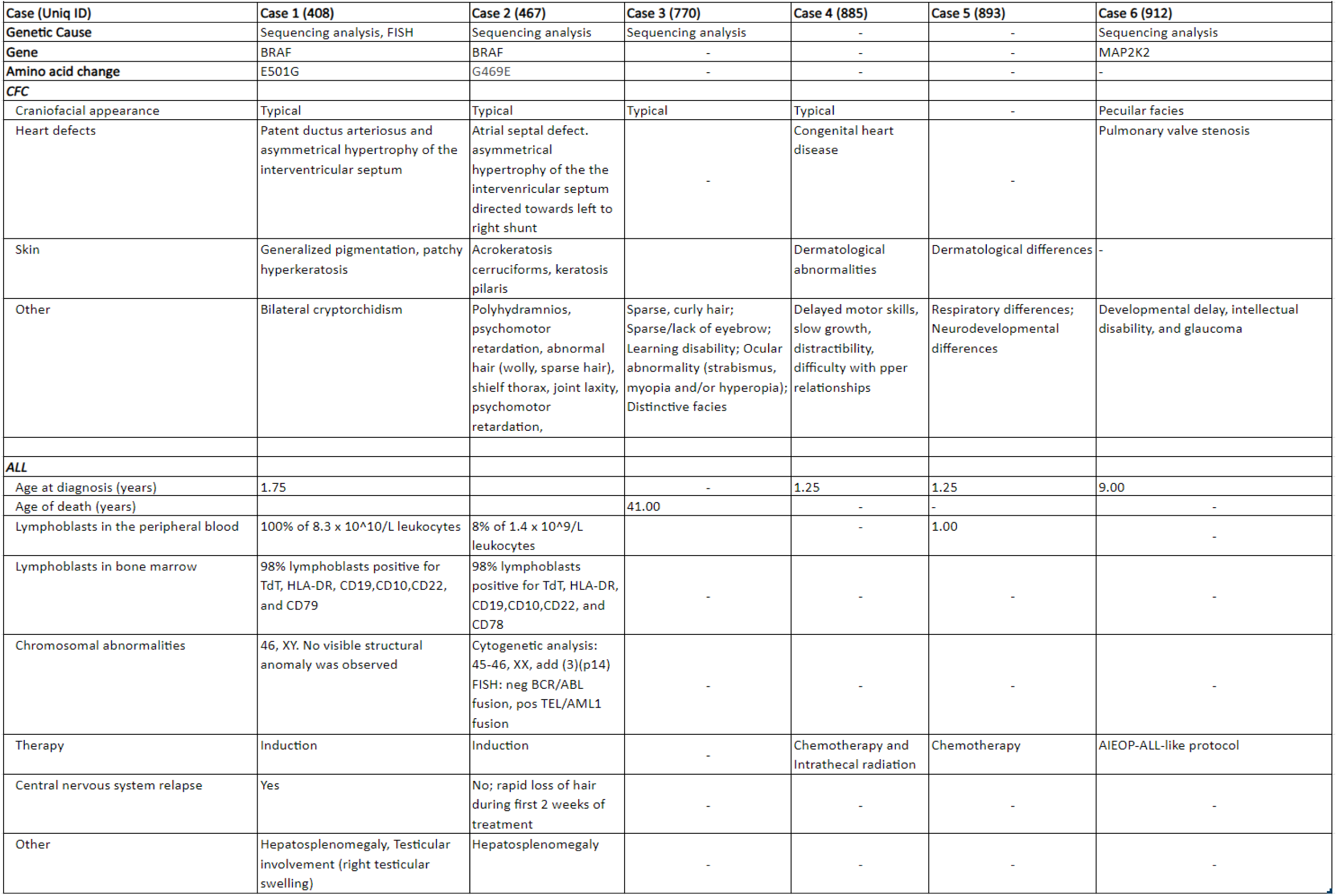
Clinical findings, presentation, and genetic cause in CFC cases with acute lymphoblastic leukemia.

### Incidence and cause-specific hazards

We aimed to understand the cumulative incidence for competing risks for cancer and death prior to cancer onset within the CFC cohort. There was a cumulative incidence, after accounting for competing risk, of 3% (95% CI:1.1-5.5) for cancer and 2% (95% CI:0.4-3.7) for death prior to cancer by age 10 (**Figure 3-A).** Therefore, 5% of CFC patients developed cancer or died by 10 years of age. The cause-specific hazard of death was 1-2% until 3 years of age and declined thereafter **(Figure 3-B).**

**Figure 3.**
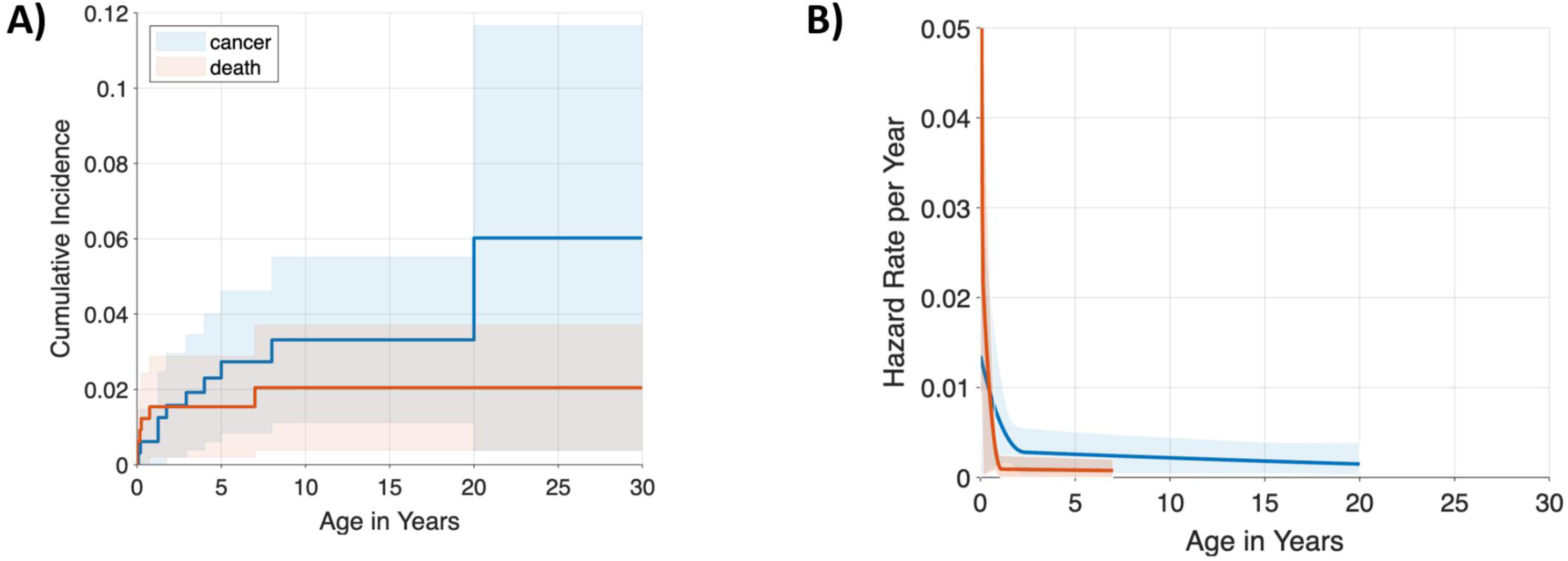
A-B. Cumulative incidence and cause-specific hazard rates by age Figure 3. (A) Cumulative incidence for cancer (blue) and cancer-free death (red) in patients with Cardiofaciocutaneous syndrome and 95% CIs (shaded regions); (B) Annual hazard rates (incidence rate per year among subjects who are still susceptible) cancer (blue) or cancer-free death (red) and 95% point-wise confidence envelopes (shaded regions) for Costello syndrome

We performed a sensitivity analysis for competing risk of cancer and death prior to cancer using tier 1 data only (P/LP variants) on patients with a reported age (n=258). The estimated cumulative incidence and cause-specific hazard was similar despite only using the subset with molecular data **(Supplementary Figure 2)**. To estimate the cancer risk associated with CFC, we calculated SIR using the six of eleven cancers that included demographic data across 4,913 person-years **(Table 3)**. All sites (SIR=4.96) and acute lymphoblastic leukemia (SIR=24.23) were significantly higher than in the general population (p<0.05). Incidence rates were stratified by attained age (0-4, 5-9,…,85+). For SIR calculations, the observed time in the cohort was matched with the appropriate attained age category to calculate expected cancers. Person-year contribution was based on when a person entered and exited. To calculate overall SIR, a sum of all the component expected cancer values were obtained to determine the overall number of expected cancers.

**Table 3.** Standardized incidence ratios for each type of cancer in CFC patients.

| Cancer type | Observed | Expected | SIR (95% CI) | Age at Event * |
| --- | --- | --- | --- | --- |
| All sites | 6 | 1.21 | **4.96(1.81-10.79)** | 3.00,4.00,6.00,9.00,12.00 |
| Acute Lymphoblastic leukemia | 4 | 0.17 | **24.23(6.52-62.03)** | 4.00, 6.00, 9.00 |
| Non-Hodgkin lymphoma | 1 | 0.06 | 15.89(0.21-88.40) | 12.00 |
| Hepatoblastoma | 1 | 0.02 | 49.57(0.65-275.80) | 3.00 |

## Discussion

This study enhances and confirms previous analyses through a comprehensive SER by creating a retrospective cohort of individuals with CFC. Our global literature cohort is currently the largest to date including 690 individuals with a clinical and/or molecular diagnosis of CFC. Despite the rarity of CFC, we were able to collect a substantial amount of CFC cases from a total of 578 case series (3 were cancers) and 112 case reports (8 were cancers). Although this is a 33% increase in case numbers from the previous systematic review in 2011, cancer cases remained similar in proportion.^12^ Since the previous systematic review, new cancer cases have been reported, including disseminated oligodendrogliallike leptomeningeal tumor ^21^and pediatric ALL.^22–24^

Exploratory analyses and descriptive statistics were conducted to understand trends in the data, finding common pediatric cancers, including ALL, most frequent in the CFC cohort. Previously reported CFC cases with cancer often provided molecular diagnoses information^6–10^, but four new cancer cases reported since the previous systematic review had no molecular diagnosis information available.^21–24^. Nonetheless, we sought to understand genotype-phenotype (cancer) correlations by examining the landscape of reported variants and correlated cancers. Six of the 11 cancers reported pathogenic variants found in the CFC cohort including *MAP2K2* p.Pro128Gln; *BRAF* p.Gly469Glu; and *MAP2K1p*.Tyr130Cys, reported in ALL, non-Hodgkin’s lymphoma, and hepatoblastoma, respectively. This molecular information is critical and can provide medical providers with an insightful clinical application tool for improved cancer risk assessment and counseling for families impacted by CFC. In addition to confirming a low cumulative incidence found in the previous study,^12^ we captured significant SIRs for all sites and ALL compared to the general population. These results suggest that cancer risk is elevated in this population, however, may be limited to early childhood.

A major limitation in this study was sparseness of data, including a modest number of cancer cases, molecular variant information, and demographics. Insufficient and/or missing data reduced our ability and statistical power to perform more in-depth analyses of genotype-phenotype associations, survival rates, and stratification by age, race/ethnicity, and other demographic factors. The reliance on published reports also introduced publication bias including possible duplicate cases, missing cases, and possible enrichment of cancer cases versus non-cancer cases. Additionally, due to the clinical overlap between RASopathy syndromes, clinical-only cases (i.e., those without a molecular diagnosis) could genetically be a RASopathy other than CFC. To address this issue, we created a tiering system to analyze and isolate molecularly confirmed CFC cases and cancer risk. Importantly, the sensitivity analysis of cumulative incidence and cause-specific hazard did not show meaningful differences between tier 1 (molecular diagnoses) and tier1 + tier 2 data (molecular and clinical diagnoses).

Our review highlights the need for further studies in this rare population to be conducted and capture the most up-to-date information regarding CFC prevalence and cancer risk. Currently, RASopathy studies are being conducted through NCI-sponsored intramural and extramural collaborations seeking to understand cancer risk throughout the lifetime of individuals with CFC. These studies will impact screening recommendations for cancer and, perhaps, better define what genotypes are most commonly associated with cancer.^4^ Together, they may improve diagnostics, clinical care management, screening guidelines, targeted therapies, and risk support for patients with RASopathies including CFC within and beyond childhood developmental stages of life.

The content of this publication does not necessarily reflect the views or policies of the Department of Health and Human Services, nor does mention of trade names, commercial products or organizations imply endorsement by the U.S. Government.

## Acknowledgements

This study was supported by the Intramural Research Program of the Division of Cancer Epidemiology and Genetics, National Cancer Institute, and National Institutes of Health. This work utilized the computational resources of the NIH HPC Beowulf cluster. (http://hpc.nih.gov)

## Additional Information

### Authors’ contributions

DS, GN, TB, AL designed and initiated framework of review, database search, and protocol. Title and abstract screening were performed by TB, JB, SB, and MA. Full text review was performed by TB, JB, SB, and MA. TB and JB performed JBI bias assessment for case reports and case series. TB, JB, AF contributed to data extraction. JB, GN, TB, PR, AP, and JK performed data cleaning. JK and AP performed variant classification. PR and JK performed data analysis. Manuscript was prepared by JB, TB, GN, and DS.

### Ethics approval and consent to participate

No use of human subjects, not applicable.

### Consent for publication

No use of human subjects, not applicable.

### Data availability

Data sharing not applicable to this article as no datasets were generated or analyzed during the current study.

### Competing interests

None.

### Funding information

The author(s) received no specific funding for this work.

## Supplementary Files

**Supplementary Methods**

**Supplementary Table 1. CFC Review Data Dictionary**

**Supplementary Table 2. CFC Final Extraction Table**

**Supplementary Table 3. Description of subsets of cases included for each analysis**

**Supplementary Table 4. Demographics**

**Supplementary Table 5. Top 3 pathogenic/likely gene variant frequency found in CFC patients**

**Supplementary Table 6. Risk of bias assessment JBI Critical Appraisal Checklist for Case Series**

**Supplementary Table 7. Risk of bias assessment JBI Critical Appraisal Checklist for Case Reports**

**Supplementary Figure 1. PRISM flowchart**

**Supplementary Figure 2. Cumulative incidence and cause-specific hazard rates by age for all tier 1 cases with pathogenic/likely pathogenic variants sensitivity analysis (n=510)**

## Notes

### Competing Interest Statement

The authors have declared no competing interest.

