## Supplementary figures and images for "A Literature Review and Pooled Case Analysis of Cardiofaciocutaneous Syndrome to Estimate Cancer Risk"

### Supplemental Figure 1

**Supplementary Figure 1.** PRISMA flowchart

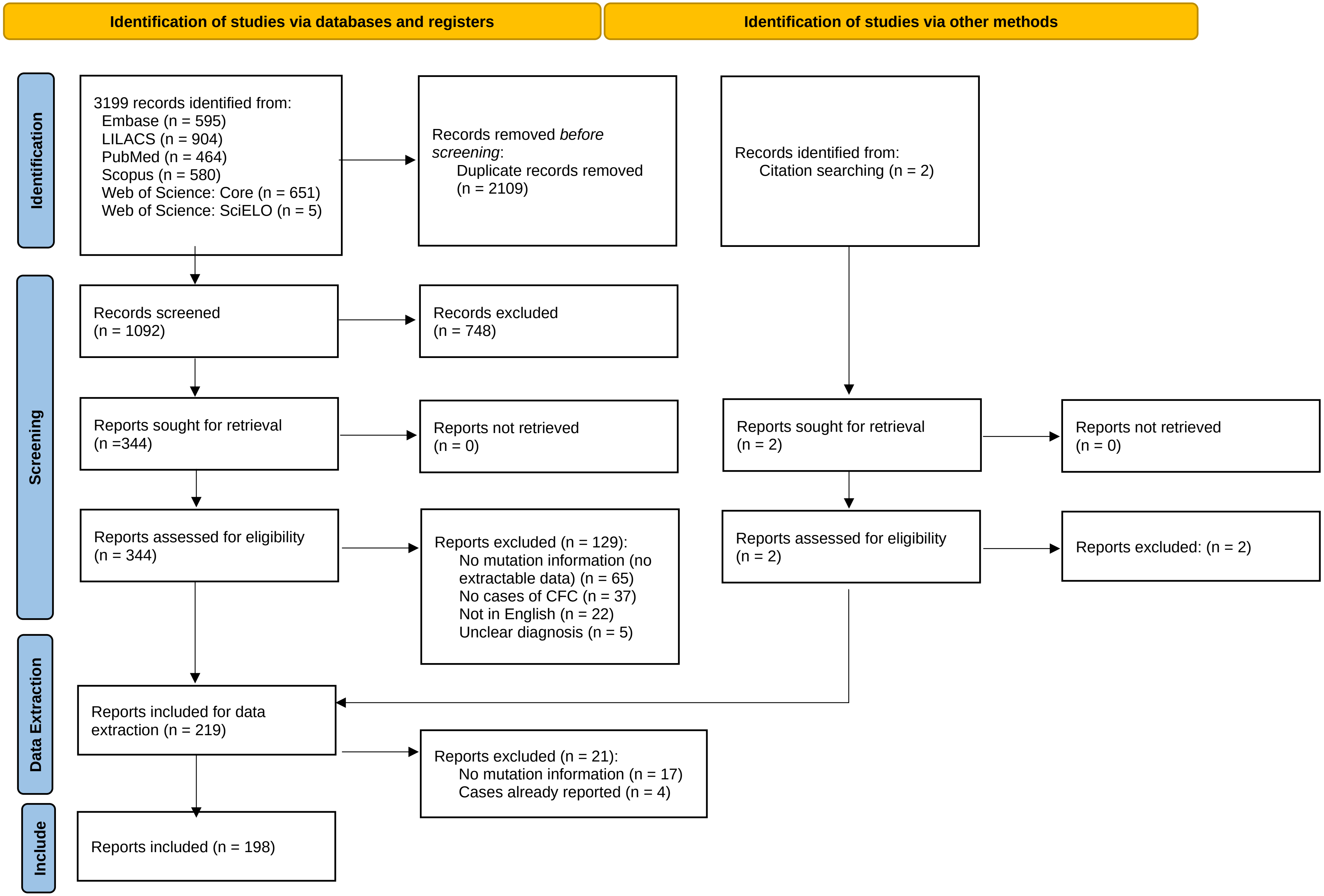
