## Supplemental Figure 2 for "A Literature Review and Pooled Case Analysis of Cardiofaciocutaneous Syndrome to Estimate Cancer Risk"

**Supplementary Figure 2A-B.** Cumulative incidence and cause-specific hazard rates by age for all tier 1 cases with pathogenic/likely pathogenic variants sensitivity analysis (n=510)

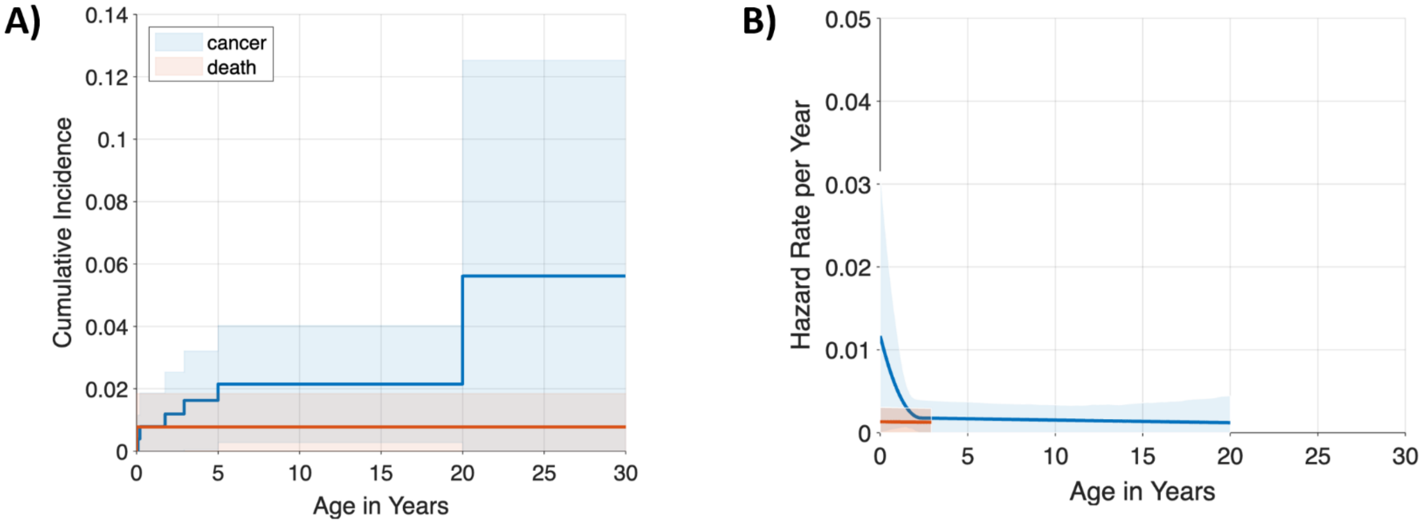


Figure 3. (A) Cumulative incidence for sensitivity analysis for cancer (blue) and cancer-free death (red) in patients with Cardiofaciocutaneous syndrome and 95% CIs (shaded regions); (B) Annual hazard rates (incidence rate per year among subjects who are still susceptible) cancer (blue) or cancer-free death (red) and 95% point-wise confidence envelopes (shaded regions) for Costello syndrome
