## Supplemental Methods for "A Literature Review and Pooled Case Analysis of Cardiofaciocutaneous Syndrome to Estimate Cancer Risk"

### Supplementary Methods

**Research questions:**

1.What is the full spectrum of tumors found in Cardiofaciocutaneous Syndrome (CFC)?
2.What are the cause-specific hazards and cumulative incidence accounting for competing risks for malignancy and death prior to cancer in CFC?
3.What is the age of presentation of these malignancies?
4.What genotypes in CFC correlate with a cancer phenotype?
5. What is the spectrum of variants found in CFC?
6. What are the Standardized incidence ratios for each cancer in CFC?

**Case definition (s):**

Cardiofaciocutaenous Syndrome: both clinical and/or molecular diagnoses will be eligible.

**Participants/population:**

Studies with patients of any age, sex/gender, race/ethnicity with a clinical and/or molecular diagnosis of CFC. Reported cases with variants of BRAF, KRAS, MAP2K1, and MAP2K2 genotypes that will be classified as pathogenic, likely pathogenic, or VUSs according to ClinVar and or InterVar guidelines for variant interpretation and classification.

**Search Strategy**

**Databases searched:**

- PubMed/MEDLINE (National Library of Medicine)
- Embase (Elsevier)
- Scopus (Elsevier)
- Web of Science: Core Collection & Scielo (Clarivate Analytics)
- Latin American and Caribbean Health Sciences Literature (LILACS)/ Pan-American Health Organization (PAHO)
- Scopus
- Google Scholar (as a supplemental search after other databases have been searched)

**Limits for search:**

- Publication date: 1980- Feb 2023

Although CFC was first named and identified in 1986, the timeline has been extended to include any potential retroactively identified cases included in literature. The diagnostic criterion set in 1986 (Reynolds, 1986) will be used to include/exclude any cases identified.

- Language: English only (originally all languages)

- Publication type: include original research articles, case reports/series, and cohort studies
- Species: Human

**Grey literature sources searched:**

- None.

**Database Search Results Table**

| Databases | Results |
|---|---|
| Embase (Elsevier) | 595 |
| LILACS (BIREME) | 904 |
| PubMed (US National Library of Medicine) | 464 |
| Scopus (Elsevier) | 580 |
| Web of Science: Core Collection (Clarivate Analytics) | 651 |
| Web of Science: SciELO (Clarivate Analytics) | 5 |
| Supplemental Citation Searching | 2 |
| **TOTAL: (all database results)** | 3,201 |
| **DUPLICATES REMOVED** | 2,109 |
| **Databases** | **Results** |
| Embase (Elsevier) | 177 |
| LILACS (BIREME) | 218 |
| PubMed (US National Library of Medicine) | 440 |
| Scopus (Elsevier) | 67 |
| Web of Science: Core Collection (Clarivate Analytics) | 189 |
| Web of Science: SciELO (Clarivate Analytics) | 1 |
| Supplemental Citation Searching | 0 |
| **TOTAL UNIQUE CITATIONS:** (after duplicates removed) | 1,092 |

### Database Searches

**Database:** PubMed/MEDLINE
Platform: US National Library of Medicine

Date Searched: February 12, 2023
Date Limits: 1980–2023
Other Limits/Filters: None

| Set | Concept | Search Strategy |
|---|---|---|
| #1 | CFC | ("cardio-cutaneous syndrome*"[Title/Abstract] OR "cardio cutaneous syndrome*"[Title/Abstract] OR "cardiofaciocutaneous syndrome*"[Title/Abstract] OR "cardio faciocutaneous syndrome*"[Title/Abstract] OR "cardio facio cutaneous syndrome*"[Title/Abstract] OR "cardio facial cutaneous syndrome*"[Title/Abstract] OR "cardio-facio-cutaneous syndrome*"[Title/Abstract] OR "neuro cardio facial cutaneous syndrome*"[Title/Abstract] OR NCFC[Title/Abstract] OR "CFC syndrome*"[Title/Abstract] OR "Cardiofaciocutaneous syndrome" [Supplementary Concept]) |
| #2 | Limits Applied: Publication Year | #1 AND (1980:2023[pdat]) |

**Notes:** The limit for publication year (1980–2023) was applied to the main search using the filters available in PubMed. The keywords were searched in the title and abstract fields (i.e., [Title/Abstract]), and the controlled vocabulary term is indicated with [Supplementary Concept]. Phrases were enclosed in quotation marks to force the searching of the exact terms in order presented. No other limits were applied to the searches.

**Database:** Embase
Platform: Elsevier
Date Searched: February 12, 2023
Date Limits: 1980–2023
Other Limits/Filters: Source: Embase and Embase Classic

| Set | Concept | Search Strategy |
|---|---|---|
| #1 | CFC | ('cardiofaciocutaneous syndrome'/exp OR 'cardio facio cutaneous syndrome'/exp OR 'cardio-cutaneous syndrome*':ti,ab OR 'cardio cutaneous syndrome*':ti,ab OR 'cardiofaciocutaneous syndrome*':ti,ab OR 'cardio faciocutaneous syndrome*':ti,ab OR |

| | | |
|---|---|---|
| | | 'cardio facio cutaneous syndrome*':ti,ab OR 'cardio facial cutaneous syndrome*':ti,ab OR 'cardio-facio-cutaneous syndrome*':ti,ab OR 'neuro cardio facial cutaneous syndrome*':ti,ab OR NCFC:ti,ab OR 'CFC syndrome*':ti,ab) |
| #2 | Limits Applied: Source | #1 AND ([embase]/lim OR [embase classic]/lim) |
| #3 | Limits Applied: Publication Year | #2 AND [1980-2023]/py |

**Notes:** The limit for publication year (1980–2023) and source (Embase or Embase Classic) were applied to the main search using the filters available. The keywords were searched in the title and abstract fields (i.e., :ti,ab), and the controlled vocabulary terms are indicated with /exp. Phrases were enclosed in quotation marks to force the searching of the exact terms in order presented. No other limits were applied to the searches.

**Database:** Scopus
Platform: Elsevier
Date Searched: February 12, 2023
Date Limits: 1980–2023
Other Limits/Filters: None

| Set | Concept | Search Strategy |
|---|---|---|
| #1 | CFC | Title-Abs-Key(("cardio-cutaneous syndrome*" OR "cardio cutaneous syndrome*" OR "cardiofaciocutaneous syndrome*" OR "cardio faciocutaneous syndrome*" OR "cardio facio cutaneous syndrome*" OR "cardio facial cutaneous syndrome*" OR "cardio-facio-cutaneous syndrome*" OR "neuro cardio facial cutaneous syndrome*" OR NCFC OR "CFC syndrome*")) |
| #2 | Limits Applied: Publication Year | #1 AND ((PUBYEAR > 1980)) |

**Database:** Web of Science: Core Collection*
Platform: Clarivate Analytics

**Date Searched:** February 12, 2023
Date Limits: 1980–2023
Other Limits/Filters: None

| Set | Concept | Search Strategy |
| --- | --- | --- |
| #1 | CFC | TS=("cardio-cutaneous syndrome*" OR "cardio cutaneous syndrome*" OR "cardiofaciocutaneous syndrome*" OR "cardio faciocutaneous syndrome*" OR "cardio facio cutaneous syndrome*" OR "cardio facial cutaneous syndrome*" OR "cardio-facio-cutaneous syndrome*" OR "neuro cardio facial cutaneous syndrome*" OR NCFC OR "CFC syndrome*") |
| #2 | Limits Applied: Publication Year | #1 AND PY=(2003-2023) |

*Science Citation Index Expanded (SCI-EXPANDED)--1900-present
Social Sciences Citation Index (SSCI)--1900-present
Conference Proceedings Citation Index – Science (CPCI-S)--1990-present
Conference Proceedings Citation Index – Social Science & Humanities (CPCI-SSH)--1990-present
Book Citation Index – Science (BKCI-S)--2005-present
Book Citation Index – Social Sciences & Humanities (BKCI-SSH)--2005-present
Emerging Sources Citation Index (ESCI)--2005-present
Current Chemical Reactions (CCR-EXPANDED)--1985-present
Index Chemicus (IC)--1993-present
**Database:** Web of Science: SciELO
Platform: Clarivate Analytics
**Date Searched:** February 12, 2023
Date Limits: 1980–2023
Other Limits/Filters: None

| Set | Concept | Search Strategy |
| --- | --- | --- |
| #1 | CFC | TS=("cardio-cutaneous syndrome*" OR "cardio cutaneous syndrome*" OR "cardiofaciocutaneous syndrome*" OR "cardio faciocutaneous syndrome*" OR "cardio facio cutaneous syndrome*" OR "cardio facial cutaneous syndrome*" OR "cardio-facio-cutaneous syndrome*" OR "neuro cardio facial cutaneous syndrome*" OR NCFC OR "CFC syndrome*") |
| #2 | Limits Applied: | #1 AND PY=(2003-2023) |

| | |
|---|---|
| | Publication Year |

**Database:** LILACS/BIREME
Platform: PAHO Virtual Health Library https://lilacs.bvsalud.org/en/
Date Searched: February 12, 2023
Date Limits: 1980–2023
Other Limits/Filters: None

| Set | Concept | Search Strategy |
|---|---|---|
| #1 | CFC 1 | Title, Abstract, Subject: ("cardio-cutaneous syndrome*" OR "cardio cutaneous syndrome*" OR "cardiofaciocutaneous syndrome*") |
| #2 | Limits Applied: Publication Year | #1 AND (year_cluster:[1980 TO 2023]) |
| #3 | CFC 2 | Title, Abstract, Subject: ("cardio faciocutaneous syndrome*" OR "cardio facio cutaneous syndrome*" OR "cardio facial cutaneous syndrome*") |
| #4 | Limits Applied: Publication Year | #3 AND (year_cluster:[1980 TO 2023]) |
| #5 | CFC 3 | Title, Abstract, Subject: ("cardio-facio-cutaneous syndrome*" OR "Neuro cardio facial cutaneous syndrome*" OR NCFC OR "CFC syndrome*") |
| #6 | Limits Applied: Publication Year | #5 AND (year_cluster:[1980 TO 2023]) |
| #7 | | #2 OR #4 OR #6 |

**Additional Methods Information:**

Data Management

For the literature searches, a biomedical librarian will use EndNote 20 (Clarivate Analytics) to collect citations, identify duplicates, and compile the unique citations for screening. All unique citations will be exported into Covidence (Veritas Health Innovation, Ltd.).

Covidence will be used to conduct the title and abstract screening and full text review steps. An online Google Form will be used to extract data from each case. These variables will be converted to a Microsoft Excel sheet to be shared with a statistician for the meta-analysis.

Inclusion criteria:

- Clinical and/ or molecular diagnosis of CFC syndrome
- Molecular information provided for BRAF, MAP2K1, MAP2K2, and KRAS genes
- Type of study designs included were case reports, case series, and cohort studies.
- English language only

Exclusion criteria:

- Other RASopathies as a primary focus (e.g., Noonan Syndrome, Legius Syndrome, Capillary Arteriovenous Malformation Syndrome, and Hereditary gingival fibromatosis, etc.)
- Animal studies
- No CFC case reported
- No diagnosis reported
- Other genotypes reported that were not BRAF, MAP2K1, MAP2K2, and KRAS
- Aggregate cases and no extractable information
- Studies in languages other than English

Risk of Bias in Individual Studies

The Johanna Briggs Institute's critical appraisal tools for case reports and case series will be used after full text review (Moola et al., 2020; Munn et al., 20) **(Supplementary File 2 and 3)**. Risk of bias for each article will be independently assessed by two reviewers (TB and JB). Both reviewers must agree on inclusion for an article's case(s) to remain in the retrospective cohort. In cases of conflict, a third reviewer (GN) will decide whether an article has inherent bias and should be excluded. All tools and processes will be piloted prior to use.

We will include from our meta-analysis any articles rated at a high risk of bias to have the most robust retrospective cohort as possible.

Data Variables

We will model our data extraction on those variables collected from Astiazaran-Symonds et al., (2023).[1]

For each included individual record, we will extract the following data variables when available. If data is unavailable, the entry will be recorded as "Missing" with a ".":

- Sex
- Ethnicity/Race/Nationality

- Age at publication in years
- Age at diagnosis of CFC in years
- Molecular Diagnosis if available
- Specific variant if available (BRAF, MAP2K1, MAK2K2, and KRAS)
- Cancer status (yes or no)
- Cancer type
- Country of origin - cancer cases only
- Benign tumor type
- Age at cancer diagnosis
- Vital status
- Age at death
- Last known age alive

Race and Ethnicity Classification

Race and ethnicities reported for each CFC case (those in parenthesis) are grouped into categories (those in bold) used by the NIH OMB categories and descriptions for race and ethnicity (Supplementary Table 1 and 4).[2]

- Asian (Asian)
- Black or African American or African descent (Non-Hispanic African American and North African)
- White (Caucasian-Cajun, Dutch, European, French, German, Italian, Middle Eastern, Non-Hispanic White, Non-Hispanic; Polish, Spanish, White; Polish, White)
- More than one race: (Brazilian, Caribbean, Non-Hispanic White and Asian, North African and Italian)
- Unknown race (Hispanic and cases with no reported race)
- Ethnicity: Hispanic or Not Hispanic

Tier-Binning Categorization for Variant Classification

Cases are categorized based on two different tier criteria as described below for all case analysis and sensitivity analysis looking at variants among cancer and not cancer cases (Supplementary Table 1).

- Tier 1: Tier 1: P/LP variants in ClinVar and/or InterVar and associated clinical details
- Tier 2: VUSs, deletion cases, cases with variant typos, and cases with clinical diagnosis only

Exploratory Analysis and Statistical Analysis

Descriptive statistics will be calculated using Microsoft Excel and table summaries for genetic variant frequencies, genetic variant frequencies with correlation to clinical phenotypes (cancer), total case counts from each article with number cancer free and number with cancer will be reported.

Lollipop plots will be generated to see the genetic landscape and frequencies of variants found in BRAF, MAP2K1, MAPK2, and KRAS among cancer-free and cancer cases.

Cause-specific hazards and cumulative incidence curves accounting for competing risks for cancer and death prior to cancer will be calculated for overall cohort and a subset of cases (tier 1) with only pathogenic/likely pathogenic variants.

We will calculate the standardized incidence ratio (SIR) comparing the incidence of cancer in cases in the literature with the expected incidence of cancer in the Surveillance, Epidemiology, and End Results (SEER) Program Database using SEER*Stat statistical software (Version 8.4.0.1). Birthdates will be estimated based on date of publication and age at publication. A SEER*Stat-ready file will be prepared using SEER*Prep and an incidence rate file will be created using SEER data for each type of cancer reported in the literature cases. SIR analyses were performed overall and stratified by birth cohort and attained age. An Excel report with the SIR results will then be created. The ratio of observed to expected incidence measures the relative incidence of cancer in this retrospective cohort will be compared to an age-matched cohort from the general population. We will then calculate 95% confidence intervals (CIs) for the incidence ratio.
