## Supplemental Table 3 for "A Literature Review and Pooled Case Analysis of Cardiofaciocutaneous Syndrome to Estimate Cancer Risk"

**Supplementary Table 3.** Description of subsets of cases included for each analysis

| **Descriptive analyses** | **No. of cases** | **Table/Figure** |
|---|---|---|
| All cases from the literature | 690 | Supp. Table 2 |
| Cases with cancer (Acute Lymphoblastic Leukemia) | 6 | Table 1 and 2, Supp. Table 2 |
| Cases with only clinical diagnosis | 150 | Supp. Table 2 |
| Cases with clinical diagnosis and molecular diagnosis (variant) available | 540 | Supp. Table 2 |
| Cases with variants classified as pathogenic or likely pathogenic | 510 | Supp. Table 2 |
| Cases with variants classified as variant of uncertain significance or likely benign | 25 | Supp. Table 2 |
| **Quantitative analyses** | | |
| Cases with sufficient clinical data for quantitative analyses | 690 | Supp. Table 2 |
| Cases who developed cancer or died | 17 | Table 1, Supp. Table 2 |
| Case who developed cancer but did not die | 8 | Table 1, Supp. Table 2 |
| Cases with tier 1 classification | 510 | Supp. Table 2 |
| Cases with tier 2 classification | 180 | Supp. Table 2 |
