## Supplemental Table 4 for "A Literature Review and Pooled Case Analysis of Cardiofaciocutaneous Syndrome to Estimate Cancer Risk"

**Supplementary Table 4.** Demographics of CFC patients

| | **Number of patients (%)** |
|---|---|
| **Sex** | |
| Female | 244 (35.4%) |
| Male | 225 (32.6%) |
| Unknown | 221 (32.0%) |
| **Age range at publication in years** | |
| 0-5 | 144 (20.9%) |
| 6-10 | 125 (18.1%) |
| 11-15 | 63 (9.1%) |
| 16-20 | 36 (5.2%) |
| ≥21 | 45 (6.5%) |
| Unknown | 277 (40.1%) |
| **Race** | |
| Black or African American or African descent | 3 (0.4%) |
| Asian | 40 (5.8%) |
| White | 103 (14.9%) |

| | |
|---|---|
| More than one race | 10 (1.4%) |
| Unknown | 534 (77.4%) |
| **Ethnicity** | |
| Hispanic or Latino* | 17 (2.5%) |
| Not Hispanic or Latino | 154 (22.3%) |
| Unknown | 519 (75.2%) |
| **Phenotype (n=690)** | |
| Cancer (All Types) | 11 (1.6%) |
| Acute lymphoblastic leukemia | 6 (0.9%) |
| Non-Hodgkin lymphoma | 2 (0.3%) |
| Rhabdomyosarcoma | 1 (0.1%) |
| Disseminated oligodendrogliallike leptomeningeal tumor | 1 (0.1%) |
| Hepatoblastoma | 1 (0.1%) |
| Cancer-free | 679 (98.4%) |

The *n* is the number of patients for each demographic characteristic , % reported out of total CFC patients (n=690).

* Cases with Hispanic or Latino ethnicity were categorized as ‘Unknown race’.
